# Imaging, Pulmonary Function, and Histopathologic Findings of Persistent Fibrosis in a Longitudinal Cohort Three Years after Severe COVID-19 Infection

**DOI:** 10.1101/2024.10.16.24315602

**Authors:** Scarlett O. Murphy, Claire F. McGroder, Mary M. Salvatore, Belinda M. D’Souza, Kathleen M. Capaccione, Anjali Saqi, Faisal Shaikh, Shannon Benesh, David Zhang, Matthew R. Baldwin, Christine Kim Garcia

**Affiliations:** Division of Pulmonary, Allergy, and Critical Care Medicine, Columbia University Irving Medical Center; New York, NY, 10032; Department of Radiology, Columbia University Irving Medical Center; New York, NY, 10032; Department of Pathology and Cell Biology, Columbia University Irving Medical Center; New York, NY, 10032; Columbia Precision Medicine Institute, Columbia University Irving Medical Center; New York, NY, 10032

## Abstract

Fibrotic-like abnormalities are present in 60% of a single-center, longitudinal, multi-ethnic cohort 3-years after severe COVID-19. They are independently associated with male sex, low BMI, shorter telomere length, increased severity of illness, and mechanical ventilation; Black race and asthma are protective. Participants with fibrotic-like abnormalities are more likely to have reduced diffusion capacity and 6-minute walk distance. Fibrotic-like abnormalities persist but modestly improve over time. Transbronchial biopsies show small airways histopathology, consistent with high prevalence of air trapping in expiration, and infrequent interstitial thickening. This study highlights the need for continued monitoring of patients with persistent fibrosis after severe COVID-19.

## INTRODUCTION

Survivors of COVID-19 can have persistent respiratory symptoms and radiographic abnormalities years after initial infection. Large cohort studies from the UK^1^ and China^2^ have reported that 11 to 36% have residual lung abnormalities following COVID-19. Radiographic imaging typically shows attenuation of inflammatory findings, including ground glass and consolidation, during the first year. However, fibrotic-like abnormalities of reticulation and traction bronchiectasis often persist, especially in patients who initially had severe or critical illness^2 3^. These residual fibrotic changes are associated with restriction and reduced diffusion capacity^2 3^. To our knowledge, there are no studies detailing histopathological correlates of persistent fibrotic changes 3-years post-COVID-19. We report longitudinal outcomes of a multi-ethnic cohort of severe COVID-19 survivors from New York City at 3-years, including radiologic, physiologic, functional, symptomatic assessments of 102 participants, and histopathological analysis of transbronchial lung biopsies for five participants with persistent fibrosis.

## METHODS

We enrolled community dwelling adults ≥21 years who were hospitalized for severe or critical COVID-19 between March 1^st^ and May 15^th^, 2020, with sampling weighted to include ∼50% requiring invasive mechanical ventilation. Patients who had participated in our prior studies at 4-months^4^ and 15-months^5^ were first approached for enrollment at 3-years. We recruited additional patients with the same inclusion criteria and sample weighting to maintain a sample size ∼100 (**Figure S1**, supplement methods). All participants completed high-resolution thoracic computed tomography (HRCT), pulmonary function testing, and symptom questionnaires. Five participants with fibrosis consented to bronchoscopy and transbronchial biopsy for histopathologic analysis.

We tested associations between clinical characteristics, genomic biomarkers and radiographic patterns using Mann-Whitney, Wilcoxen signed Rank, McNemar’s and Fischer exact tests. We calculated Spearman’s rank correlation coefficients between HRCT scores and 3-year clinical measurements. We included statistically significant univariate associations that were biologically plausible in adjusted analyses. To avoid confounding and to minimize overparameterization, we generated covariate balanced propensity scores (CBPS) that included possible confounding variables. We used generalized additive logistic models (GAMs) to assess for potential non-linear associations between risk factors and fibrosis. We estimated odds ratios via logistic regression if there was no evidence of non-linearity. For risk factors with non-linear associations, we estimated tertile-specific odds ratios.

## RESULTS

Three years after acute COVID-19 hospitalization, we enrolled 102 participants with a mean(SD) age of 60(12) years; 55(54%) were male, 62(61%) were non-White, and 50(49%) required mechanical ventilation (**Table S1**). Over 70% previously participated in our 4-month and/or 15-month post-COVID studies (**Figure S1**).^5^ While there was no difference in demographics or clinical variables between those who did or did not follow up after the 15-month study, the 27 participants who were newly recruited to participate in the 3-year study were slightly older and reported decreased physical activity (**Table S2** and **Table S3**). More than 67% had radiographic abnormalities, including 61% with fibrotic-like abnormalities, characterized by reticulations and traction bronchiectasis (**Figure 1, Tables S4** and **S5**).

**Figure 1.**
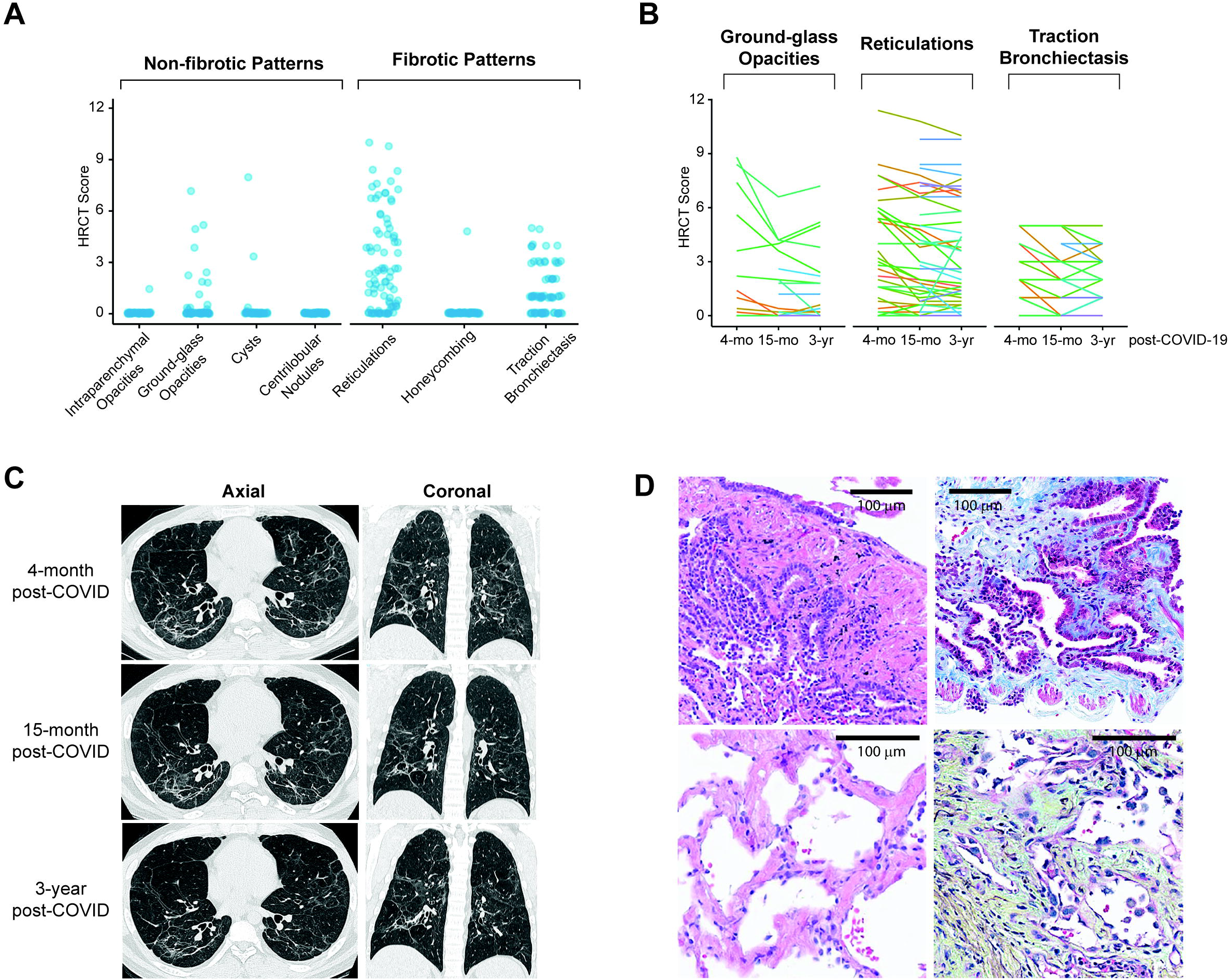
**A**. High resolution Computed Tomography (HRCT) chest scores for 102 post-COVID-19 survivors at 3 years. The patterns are segregated into non-fibrotic and fibrotic patterns. **B**. Change in individual CT pattern scores for 75 individuals 3-years after COVID-19 who at least one prior scan. This includes 41 participants with scans at all time points (4-month, 15-month, 3-year), 72 individuals with scans for only the last two time points, and 3 individuals with scans for only the first and last time points. Each line represents an individual. **C**. Representative example of one study participant with persistent radiographic abnormalities. HRCT reticulation scores at 4-month, 1-, and 3-year were 8.4, 7.8, and 6.8, respectively. Traction bronchiectasis scores at 4-month, 1-, and 3-year were 5, 3, and 4, respectively. **D**. Lung transbronchial biopsies of two participants whose fibrosis scores were in the top quartile. The top row shows the lung histopathology of the same participant whose HRCT scan is shown in **C**. The lung shows evidence of small airways disease with peribronchiolar metaplasia; H&E (left) and Masson trichome (right) stained slides are shown. The bottom row shows the lung histopathology of another participant (HRCT scans shown in **Figure S2** panel E) with alveolar interstitial thickening by fibromyxoid tissue and type 2 cell hyperplasia; H&E (left) and Movat (right) stained slides are shown. Bars represent 100 microns.

In unadjusted analyses, participants with fibrotic-like abnormalities were more often male, had lower BMI, shorter telomere length, higher admission sequential organ failure assessment (SOFA) scores, longer hospitalization, and more frequently required mechanical ventilation. Black race and patient-reported asthma were negatively associated with fibrosis (**Table S1**). In adjusted analyses, male sex, lower BMI, shorter telomere length, SOFA score, and days of mechanical ventilation were independently associated with increased odds of fibrotic-like abnormalities (**Table 1**), consistent with our 4-months^4^ and 15-month^5^ studies. Black race and self-reported asthma remained protective, a novel finding.

**Table 1.** Adjusted Associations between Clinical Variables and Fibrotic-like Radiographic Abnormalities at 3 Years Post-COVID.

| Variable | Odds Ratio | 95% CI | P-Value |
| --- | --- | --- | --- |
| Age at hospital admission |  |  | 0.18 |
| Tertile 1: 22-50 (ref) | 1.00 |  |  |
| Tertile 2: 51-61 | 0.95 | (0.35, 2.6) |  |
| Tertile 3: 62-78 | 2.19 | (0.77, 6.19) |  |
| Male Sex | 3.52 | (1.4, 8.85) | **0.007** |
| Age-Adjusted Telomere Length Percentile† | 0.82 | (0.66, 1) | **0.025** |
| BMI on Admission | 0.94 | (0.88, 1) | **0.044** |
| Ventilator Days | 1.08 | (1.04, 1.12) | **<0.001** |
| SOFA Score on Admission |  |  | **<0.001\*** |
| Tertile 1: 0-2 (ref) | 1.00 |  |  |
| Tertile 2: 3-4 | 1.47 | (1.54, 12.34) |  |
| Tertile 3: 5-11 | 2.54 | (3.75, 43.29) |  |
| Black or African American Race‡ | 0.37 | (0.14, 0.98) | **0.045** |
| Current or Former Smoker | 0.64 | (0.26, 1.59) | 0.34 |
| History of Asthma | 0.30 | (0.09, 0.95) | **0.041** |

Given that the study sample size would not support a single multivariable model, individual models were created for each independent variable listed in the table. Covariate balanced propensity scores specific to each independent variable were generating from the other potentially confounding covariables, including (age, sex, race, BMI, admission SOFA score, telomere length percentile, ventilator days, history of asthma, and smoking history). Abbreviations: OR, odds ratio; CI, confidence interval; BMI, body mass index; SOFA, sequential organ failure assessment.
\*P-for-trend between tertiles
†per 10% increase in telomere length
‡as compared to White, Asian, or Other Race

Fibrotic-like abnormalities were associated with lower diffusion capacity (DLCO), lower 6-minute walk distance (6MWD), lower physical activity, slower gait-speed, and higher prevalence of pre-frailty (**Table 2**). The 6MWD, but not the other markers of frailty, were associated with fibrotic-like abnormalities after adjusting for severity of illness. Reticulation and traction bronchiectasis scores were weakly correlated with DLCO and dyspnea (**Table S6**).

**Table 2.** Associations of pulmonary function, physical function, and symptoms, with fibrotic-like abnormalities at 3 years post-COVID-19.

|  | Total,<br>(N=102) | Fibrotic-Like<br>Abnormalities*<br>(N=62) | Normal or Non-fibrotic<br>Abnormality<br>(N=40) | P-value† | Adjusted<br>P-value† |
| --- | --- | --- | --- | --- | --- |
| FVC %Predicted, mean(SD)‡ | 87.5 (17.1) | 86.6 (18.2) | 87.5 (17.1) | 0.80 |  |
| Reduced FVC, N(%)‡ | 27 (26.7%) | 16 (25.8%) | 11 (27.5%) | 0.79 |  |
| DLCO %Predicted, mean(SD)‡ | 81.2 (20.2) | 77.7 (20.2) | 86.8 (19.2) | **0.028** |  |
| Reduced DLCO, N(%)‡§ | 41 (40.6%) | 32 (51.6%) | 9 (22.5%) | **0.004** |  |
| 6MWD %Predicted, mean(SD)¶ | 72.8 (18.4) | 69.8 (17.7%) | 77.3 (18.7) | **0.049** | **0.018** |
| Reduced 6MWD, N(%)¶# | 60 (61.9%) | 42 (72.4%) | 18 (46.2%) | **0.009** |  |
| Weak Grip, N(%)♣ | 41 (40.2%) | 26 (41.9%) | 15 (37.5%) | 0.66 |  |
| Decreased Activity, N(%)♣ | 10 (9.8%) | 9 (14.5%) | 1 (2.5%) | **0.046** | 0.24 |
| Exhaustion, N(%)♣ | 32 (31.4%) | 22 (35.5%) | 10 (25.6%) | 0.26 |  |
| Slow 4.57 M Walk Speed, N(%)♣ | 29 (28.4%) | 23 (37.1%) | 6 (15.0%) | **0.016** | 0.46 |
| Weight Loss > 10 lbs, N(%)♣ | 19 (18.6%) | 11 (17.7%) | 8 (20.0%) | 0.78 |  |
| Prefrail, N(%)♥ | 60 (58.8%) | 42 (67.7%) | 18 (45.0%) | **0.023** | 0.51 |
| Frail, N(%)♥ | 12 (11.8%) | 8 (12.9%) | 4 (10.0%) | 0.76 |  |
| Frail or Prefrail, N(%) | 72 (70.6%) | 50 (80.6%) | 22 (55.0%) | **0.006** | 0.65 |
| UCSD SOBQ Score, median (IQR) | 30 (9-55) | 36.5 (13-58) | 25.5 (5.5-48) | 0.12 |  |
| MMRC Dyspnea Score, mean (SD) | 1.2 (1.1) | 1.2 (1.1) | 1.2 (1.2) | 0.94 |  |
| Cough Severity ≥30, N(%) | 38 (37.2%) | 24 (38.7%) | 14 (35.0%) | 0.71 |  |

Abbreviations: FVC, forced vital capacity; FEV1, forced expiratory volume in 1 second; DLCO, diffusion capacity for carbon monoxide; 6MWD, six-minute walk distance; VAS, visual analogue scale from 0 - 100.
\*Fibrotic-like abnormalities include reticulations, traction bronchiectasis, or honeycombing
†Exact McNemar's test for comparison of binary outcomes and Wilcoxon signed rank test for comparison of continuous outcomes. The adjusted p-value was generated using a covariate balanced propensity model including the indicated variable, age, sex, SOFA score at admission, maximum level of oxygen support during hospitalization, and the hospital length of stay.
‡Data missing for one individual (N=101 for full group, N=39 for those with non-fibrotic abnormalities)
§Per 2017 ATS/ERS guidelines
¶Data missing for 4 individuals (N=97 for full group, N=58 for those with fibrotic changes, N=39 for those with non-fibrotic changes)

Among participants with thoracic CT abnormalities who also participated at 4-months or 15-months, fibrotic-like abnormalities improved over time, with decreasing reticulation scores (**Table S7**). For 56 participants with imaging at 4-months and 3-years, 29% showed improvement and the rest had stable findings (**Table S8**). For 77 participants with imaging at 15-months and 3-years, 9% showed improvement and the rest had stable findings. None had worsening fibrosis. Among those who were evaluated at 15-months and 3-years, there was more air trapping at 3-years (88.7% vs. 48.6%, p<0.001, **Table S9**). Air trapping was partially explained by asthma but was not associated with fibrotic-like abnormalities (**Table S10**). Participants had higher cough scores at 3-years than at 15-months, but cough was not associated with air trapping or fibrosis (**Tables S10 and S11**).

Five participants with fibrosis scores in the top quartile (**Table S12, Figure S2**) agreed to undergo bronchoscopy with transbronchial lung biopsy (**Figure 1**). Evidence of small airways disease, primarily peribronchiolar metaplasia, was found in four of four biopsies containing small airways. All five biopsies contained alveoli but only one showed increased interstitial thickness and fibrosis.

## DISCUSSION

In this single-center, New York City-based, Black and Hispanic-predominant cohort of adult COVID-19 survivors 3-years from the initial wave, we find a 60% prevalence of fibrotic-like abnormalities, which is associated with reduced DLCO and 6MWD. We report the novel findings that Black race and asthma are protective of post-COVID fibrosis. Histopathological analyses show predominantly small airways disease, which is consistent with the high prevalence of air trapping on expiratory imaging, and rare interstitial thickening.

Similar to prior studies, we find that greater severity of initial illness^1-3 6^, duration of hospitalization or ventilatory support^3^, and shorter telomere lengths^7^ are independent risk factors of post-COVID fibrosis. We identify male sex as a risk factor for post-COVID fibrosis, consistent with its risk for Idiopathic Pulmonary Fibrosis (IPF). Those with lower BMI are more likely to survive COVID-19 acute respiratory distress syndrome (ARDS)^8^, and may be at increased risk of post-COVID fibrosis. People with mild or well-controlled asthma do not have an increased risk of death or COVID-19 hospitalization^9^, and here we find asthma is associated with less post-COVID fibrosis. Consistent with a report of increased air trapping within the first 6 months following COVID-19^10^, we find a high prevalence of small airways disease (89%) that is apparent in biopsy specimens.

The 60% prevalence of fibrotic-like imaging abnormalities at 3-years likely reflects the high numbers of patients who underwent mechanical ventilation. In comparison, the Wuhan, China cohort had a prevalence of fibrotic abnormalities of 47% among those requiring high-flow nasal oxygen or mechanical ventilation.^2^ While we observed normalization of pulmonary function and 6MWD at 15-months,^5^ we now find reduced DLCO and 6MWD are associated with fibrotic-like abnormalities.

Limitations of our study include the 30% attrition from the 15-month study and lower physical activity of newly recruited participants, despite having similar demographics and initial severity of acute COVID-19 illness.

In summary, we observe a high prevalence of persistent fibrotic-like abnormalities that are associated with pulmonary degradations among survivors of severe COVID-19 at 3-years. Thus, there is an ongoing urgency for continued monitoring of survivors of severe COVID-19.

## Data Availability

All data produced in the present study are available upon reasonable request to the authors

## Acknowledgements

The authors gratefully acknowledge the contributions of the COVID-19 survivors, clinical research coordinators William Quigley and Hector Himede, and technical expertise of Mason Amelotte and Lesley Vickers.

