## Supplemental Materials for "Imaging, Pulmonary Function, and Histopathologic Findings of Persistent Fibrosis in a Longitudinal Cohort Three Years after Severe COVID-19 Infection"

**Supplemental Methods**

**Table S1.** Demographic and clinical features of severe COVID-19 Survivors 3-years from initial Illness

**Table S2.** Demographic and clinical characteristics of participants of the 15-month post-COVID study who did or did not participate in the 3-year post-COVID study

**Table S3.** Demographic and Clinical characteristics of participants of the 3-year post-COVID study who had previously participated in the 15-month post-COVID study and those who were newly recruited to the 3-year study

**Table S4.** Prevalence of HRCT abnormalities 3-years post-COVID-19

**Table S5.** Semi-quantitative HRCT scores of participants

**Table S6.** Spearman correlations between HRCT scores at 3-years post-COVID with measures of pulmonary function, frailty, grip strength and dyspnea

**Table S7.** Pairwise comparisons of participants with HRCT scans with non-zero HRCT scores

**Table S8.** Qualitative assessment of HRCT changes after COVID-19

**Table S9.** Quantification of imaging findings, pulmonary function, indices of frailty and symptom scores for all participants and for the subset of participants with follow-up at both 15-months and 3-years after COVID-19

**Table S10.** Associations with air trapping on imaging studies at 3-year post-COVID-19

**Table S11.** Number of participants with and without fibrotic-like abnormalities on their 3-year HRCT chest scans who have self-reported post-COVID symptoms

**Table S12**. Characteristics of five participants who underwent bronchoscopy with transbronchial biopsy

**Figure S1.** Recruitment Flow Diagram

**Figure S2.** HRCT Findings of Five Participants who Underwent Bronchoscopy with Transbronchial Lung Biopsy

**Supplemental Methods**

*Study Design and Participants*

We conducted a single-center prospective study of adults aged 21 years and older hospitalized between March 1, 2020 and May 15, 2020 with positive SARS-CoV-2 RT-PCR nasopharyngeal swab. All patients required oxygen during their hospitalization, fulfilling criteria for severe disease^1^ or WHO severity scores^2^ of 4 (hospitalized patients requiring supplemental oxygen via nasal cannula or mask), 5 (hospitalized patients requiring high-flow nasal cannula or non-invasive mechanical ventilation), or 6 or 7 (hospitalized patients requiring mechanical ventilation, extracorporeal membrane oxygenation (ECMO), including those who also required vasopressors and renal replacement therapy). Patients were included if they were living independently in the community prior to hospitalization. Patients with a history of interstitial lung disease or lung transplantation were excluded. The study was approved by the Columbia University Irving Medical Center (CUIMC) Institutional Review Board (AAAT5605). Patients signed written consent forms; Spanish language forms and interpreter services were available.

We contacted and enrolled prospective participants meeting eligibility criteria based on their admission date by telephone, with sampling weighted to include approximately 50% survivors with WHO severity grade severity grade 6 or 7 (required mechanically ventilation). Individuals who had previously participated in either the 4- and 15-month post-COVID research study were first contacted for enrollment. Additional individuals were enrolled using the same inclusion criteria to increase the study number to ~100 participants (**Figure S1**).

*Electronic Medical Record Measurements*

Clinical data was extracted from the New York Presbyterian-CUIMC EPIC electronic medical record. Patient data included admission vital signs, ventilator setting and flowsheet data, laboratory tests, procedures, administered medications, and procedures. We prospectively assessed comorbid conditions from patients at the study visit. Clinical and laboratory values from the first 24 hours of admission were used to calculate a SOFA score^3^. Glasgow coma scale was missing for most patients, so a score of 15 was imputed to generate the SOFA score.

*Chest Computed Tomography*

All participants underwent a non-contrast high resolution CT skin at full inspiration and on exhalation using a GE-Revolution 256 ES CT Scanner. CT scans were analyzed with two thoracic radiologists (MMS, BMD) using a semi-quantitative method validated by ARDSnet^4^ previously utilized for the 4-month and 15-month post-COVID studies^5 6^. All follow-up scans for participants were examined by two thoracic radiologists together and consensus scores were generated. Images were analyzed for the presence of reticulation, honeycombing, ground glass opacities, traction bronchiectasis, diffuse centrilobular nodules, non-emphysematous cysts, and intraparenchymal opacities. Abnormalities affecting >5% of a lung slice quadrant were scored; those affecting <5% of a lung slice quadrant were not scored. Traction bronchiectasis, reticulations, and honeycombing were categorized as fibrotic; other abnormalities were categorized as non-fibrotic. For patients with serial scans, radiologists subjectively scored the series as demonstrating progression, regression, or stability. The radiologists noted the presence or absence of air trapping after review of both the inspiratory and expiratory scans.

*Clinical Measurements*

Pulmonary function testing with spirometry and DLCO measurements were performed using an EasyOne spirometer and interpreted based on established guidelines.^7^ Cough and dyspnea were assessed utilizing the 100 mm cough visual analogue scale, modified medical research council dyspnea score and the UCSD shortness of breath questionnaire. Six-minute walk distance was obtained and analyzed utilizing establish guidelines.^8^ A Columbia University Biobank post-COVID symptom survey was used to assess persistent post-COVID symptoms.

*Frailty Measurements*

We measured five Fried frailty domains: gait-speed using a 4.57 m walk test^9^, grip-strength using a hand dynameter^9^, weight loss, low physical activity using the Duke Activity Status Index Score,^10 11^ and exhaustion using a validated questionnaire^9^. Weight loss was defined as >10lb. decrease from hospitalization admission weight to follow-up weight. Participants were assigned a point for each frailty domain they met. Consistent with Cardiovascular Health Study methodology^9^, patients were categorized as prefrail if 1 or 2 domains were abnormal and were categorized as frail if ≥3 domains were abnormal.

*Bronchoscopy with Transbronchial Lung Biopsy*

Patients with the highest quartile of reticulation (>4.2) and traction bronchiectasis (>1.0) scores who did not have a safety contraindication (defined as heart failure or reduced RV systolic function on echo, need for hemodialysis, age ≥70) were invited to undergo bronchoscopic lung biopsy under conscious sedation using fentanyl and midazolam. Between 10-16 biopsies were taken from regions of the lungs that appeared to have the highest burdens of reticulations on CT scans. Biopsy samples fixed in formalin were analyzed by pathology (AS).

*Genomic Measurements*

Blood was drawn from each participant at each follow-up visit (4 months, 15-months, and 3 years). DNA was isolated from blood leukocytes utilizing the Gentra Puregene Blood kit (Qiagen, Valencia CA). Leukocyte telomere length (LTL) was measured utilizing a quantitative PCR assay with the Rotogene rtPCR system (Qiagen).^12^ The LTL was expressed as a log-transformed ratio of telomere to single-copy gene [ln(T/S)] and this value was compared to LTL from normal control subjects (n=201 unrelated multiethnic individuals from Dallas, TX, ranging in age from 19 to 89 years) to estimate an age-adjusted LTL percentile. The *MUC5B* risk allele rs35705950 genotype was determined by Sanger sequencing.

*Missing Data*

The Glasgow Coma Score, a component of the SOFA score, was missing for nearly all non-ICU patients, so we imputed a score of 15 for these participants based on previous literature^13^. Hospital admission weight and BMI data were missing for one individual with fibrotic changes. Administration of steroids and tocilizumab data were missing for one individual without fibrotic changes. Ventilator days were missing for 3 of 50 mechanically ventilated patients, two who had fibrosis and one who did not. One individual was not able to complete PFTs at the 3-year visit; four individuals were unable to complete six-minute walk testing, including three with fibrosis and one without fibrosis. Patients with missing data were omitted from univariable analyses. For adjusted analyses, patients with any missing data were excluded such that the analysis was run on data from 96 participants at year 3 with zero missingness.

*Statistical Analyses*

In adjusted analyses, to control for confounding while minimizing model overparameterization, we generated covariate balance propensity scores (CBPS) using the CBPS package in R by regressing potential confounders on the independent variable of interest. The CBPS included age, sex, race, BMI, admission SOFA score, telomere length percentile, ventilator days, history of asthma, and smoking history. Generalized additive logistic models (GAMs) were generated for each independent variable to assess for non-linearity; in the GAM analyses the CBPS was considered balanced if the spearman correlation coefficient between the covariable and variable of interest was <[0.3]. Odds ratios were generated via logistic regression if there was no evidence of non-linearity in the GAM analyses. Age and SOFA showed evidence of non-linearity; hence values were divided into tertiles to generate tertile specific odds ratios and a P-for-trend is reported to test the significance of the relationship between tertiles (see Table 1).

**Table S1. Demographic and clinical features of severe COVID-19 Survivors 3-years from initial Illness**

|  | **Total** | **Fibrotic-Like Patterns** | **Normal CT Scan or Non-Fibrotic Change** | **P-value¶** |
| --- | --- | --- | --- | --- |
| Number | 102 | 62 (60.8%) | 40 (39.2%) |  |
| **DEMOGRAPHICS** |  |  |  |  |
| Age at Hospital Admission, mean (SD) | 55.9 (12.3) | 57.2 (13.0) | 54.0 (10.9) | 0.20 |
| Age at Visit | 60.0 (12.3) | 60.2 (12.9) | 57.0 (11.1) | 0.42 |
| Male | 55 (53.9%) | 43 (69.4%) | 12 (30.0%) | **<0.001** |
| Race |  |  |  | 0.16 |
| White | 40 (39.2%) | 25 (40.3%) | 15 (37.5%) | 0.78 |
| Black | 25 (24.5%) | 11 (17.7%) | 14 (35.0%) | **0.048** |
| Asian | 1 (1.0%) | 1 (1.6%) | 0 (0%) | 0.61 |
| Other | 36 (25.3%) | 25 (40.3%) | 11 (27.5%) | 0.19 |
| Hispanic Ethnicity | 70 (68.6%) | 46 (74.2%) | 24 (60%) | 0.13 |
| BMI on Admission* | 32.6 (7.4) | 31.4 (6.2) | 34.5 (8.6) | **0.04** |
| BMI at Visit | 33.3 (8.5) | 32.1 (6.6) | 35.4 (10.6) | 0.06 |
| **GENOMIC FACTORS** |  |  |  |  |
| Leukocyte Telomere Length Percentile, Median (IQR) | 50 (46-66) | 50 (37-50) | 50.5 (49-83) | **0.02** |
| LTL <10th percentile | 11 (10.8%) | 7 (11.3%) | 4 (10.0%) | 0.56 |
| *MUC5B* Heterozygous | 5 (4.9%) | 1 (1.6%) | 4 (10.0%) | 0.07 |
| *MUC5B* Homozygous | 2 (2.0%) | 2 (3.2%) | 0 (0.0%) | 0.52 |
| **COMORBIDITIES** |  |  |  |  |
| Hypertension | 61 (59.8%) | 26 (41.9%) | 25 (62.5%) | 0.66 |
| Diabetes | 29 (28.4%) | 17 (27.4%) | 12 (30.0%) | 0.78 |
| COPD | 5 (4.9%) | 3 (4.8%) | 2 (5.0%) | 1.0 |
| Asthma | 22 (21.6%) | 6 (9.7%) | 16 (40.0%) | **<0.001** |
| Heart Disease | 4 (3.92%) | 2 (3.2%) | 2 (5.0%) | 0.64 |
| Former Smoker | 37 (26.3%) | 18 (29.0%) | 19 (47.5%) | 0.06 |
| Active Smoker | 2 (2.0%) | 0 (0.0%) | 2 (5.0%) | 0.15 |
| CLINICAL FACTORS |  |  |  |  |
| Admission SOFA, mean (SD) | 3.7 (2.3) | 4.4 (2.1) | 2.6 (2.0) | **<0.001** |
| Received Steroids† | 47 (46.1%) | 38 (61.3%) | 11 (28.2%) | **0.01** |
| Received Anti IL-6R Blocker† | 23 (22.7%) | 20 (32.3%) | 3 (7.7%) | **0.003** |
| Maximum Oxygen Requirement |  |  |  | **<0.001** |
| Nasal Canula | 32 (31.4%) | 9 (14.5%) | 23 (57.5%) | **<0.001** |
| Non-Rebreather | 18 (17.6%) | 10 (16.1%) | 8 (20.0%) | 0.61 |
| NIPPV or HFNC | 2 (2.0%) | 2 (3.2%) | 0 (0.0%) | 0.52 |
| Mechanical Ventilation | 50 (49.0%) | 41 (66.1%) | 9 (22.5%) | **<0.001** |
| Ventilator Days, Median (IQR)‡ | 35 (17-46) | 39 (20-49) | 12 (9-15) | **0.002** |
| Hospital Days, Median (IQR) | 22 (7-46) | 38 (21-55) | 7 (4-16) | **<0.001** |
| **OUTCOMES** |  |  |  |  |
| FVC, % Predicted, mean (SD)† | 87.5 (17.1) | 86.6 (18.3) | 87.5 (17.1) | 0.85 |
| Reduced FVC at 3 years† | 27 (26.7%) | 16 (25.8%) | 11 (37.9%) | 0.79 |
| DLCO, % Predicted, mean (SD)† | 81.2 (20.2) | 77.7 (20.2) | 86.8 (19.2) | **0.03** |
| Reduced DLCO at 3 years† | 41 (40.6%) | 32 (51.6%) | 9 (23.1%) | **0.003** |
| 6MWD, % Predicted, mean (SD)§ | 72.8 (18.4) | 69.8 (17.7) | 77.3 (18.7) | 0.05 |
| Reduced 6MWD at 3 Years§ | 60 (61.9%) | 42 (72.4%) | 18 (46.2%) | **0.01** |
| Change in BMI from admission, median (IQR)* | 0.7 (-.9-2.3) | 0.4 (-.9-2.4) | 0.8 (-.8-2.1) | 0.91 |
| Cough VAS, median (IQR) | 16 (0-50) | 18 (0-50) | 16 (0-48) | 0.51 |
| Cough VAS >30 | 38 (37%) | 24 (39%) | 14 (35%) | 0.71 |

*Data missing for one individual (N=101 for full group, N=61 for those with fibrotic changes.)

†Data missing for one individual (N=101 for full group, N=39 for those with non-fibrotic changes.)

‡Data missing for three individuals (N=47 for those undergoing mechanical ventilation, N=39 with fibrotic changes, and N=8 with non-fibrotic changes.

§Data missing for 4 individuals (N=97 for full group; N=58 for those with fibrotic changes, N=39 for those with non-fibrotic changes.)

¶Associations examined using Chi-square, ANOVA or Kruskall-Wallis where appropriate

Abbreviations: COPD, Chronic Obstructive Pulmonary Disease; IL-6R, interleukin 6 receptor; NIPPV, non-invasive positive pressure ventilation; HFNC, high-flow nasal cannula, MV, mechanical ventilation; ECMO, extracorporeal membrane oxygenation; FVC, forced vital capacity; DLCO, diffusion capacity for carbon monoxide; 6MWD, 6- minute walk distance.

**Table S2. Demographic and clinical characteristics of participants of the 15-month post-COVID study who did or did not participate in the 3-year post-COVID study.**

|  | **15-month Participants without Follow-Up (N=32)** | **15-month Participants with Follow-Up (N=72)** | **p-value** |
| --- | --- | --- | --- |
| DEMOGRAPHICS |  |  |  |
| Age, mean (SD) | 55.9 (9.3) | 53.8 (11.8) | 0.39 |
| Male (%) | 20 (62.5%) | 41 (56.9%) | 0.60 |
| Hispanic Ethnicity | 20 (62.5%) | 42 (58.3%) | 0.69 |
| Race |  |  |  |
| White | 10 (31.2%) | 20 (27.8%) | 0.72 |
| Black | 6 (18.8%) | 22 (30.6%) | 0.21 |
| Asian | 1 (3.1%) | 3 (4.2%) | 0.80 |
| Other | 15 (46.9%) | 27 (37.5%) | 0.37 |
| BMI on admission, mean (SD) | 32.4 (8.2) | 21.9 (6.1) | 0.71 |
| BMI at year 1, mean (SD) | 34.6 (7.7) | 35.6 (21.3) | 0.80 |
| GENOMIC FACTORS |  |  |  |
| Leukocyte Telomere Length Percentile, Mean (SD) | 41.1 (17.2) | 41.4 (23.7) | 0.95 |
| LTL <10th percentile, N(%) | 2 (6.2%) | 10 (13.9%) | 0.26 |
| MUC5B Heterozygote, N(%) | 2 (6.7%) | 5 (6.9%) | 0.90 |
| MUC5B Homozygote, N(%) | 0 (0%) | 1 (1.4%) | 0.50 |
| COMORBIDITIES |  |  |  |
| Hypertension | 16 (50.0%) | 34 (47.2%) | 0.79 |
| Diabetes | 6 (18.8%) | 6 (18.8%) | 0.69 |
| COPD | 3 (9.4%) | 1 (1.4%) | 0.05 |
| Asthma | 6 (18.8%) | 18 (25.0%) | 0.46 |
| Heart Disease | 1 (3.1%) | 3 (4.2%) | 0.80 |
| Former Smoker | 13 (40.6%) | 28 (37.5%) | 0.76 |
| Active Smoker | 1 (3.1%) | 2 (2.8%) | 0.92 |
| CLINICAL FACTORS |  |  |  |
| Admission SOFA, median (IQR) | 3 (2-5) | 3 (2-5) | 0.51 |
| Received Steroids | 13 (40.6%) | 30 (41.7%) | 0.92 |
| Received Anti IL-6R Blocker* | 8 (25.0%) | 18 (25.3%) | 0.97 |
| Maximum Oxygen Requirement |  |  |  |
| Nasal Canula | 11 (34.4%) | 20 (27.8%) | 0.50 |
| NIPPV or HFNC | 1 (3.1%) | 1 (1.4%) | 0.55 |
| Mechanical Ventilation N (%) | 14 (43.8%) | 39 (54.2%) | 0.33 |
| Ventilator Days, Median (IQR)† | 28 (11-55) | 36 (19-46) | 0.24 |
| Hospital Days, Median (IQR)† | 16 (8-34) | 30 (9-48.5) | 0.25 |

*Data missing from one participant with follow up (N=71).

†Data included for 14 participants without follow and for 39 with follow up.

**Table S3. Demographic and Clinical characteristics of participants of the 3-year post-COVID study who had previously participated in the 15-month post-COVID study and those who were newly recruited to the 3-year study.**

|  | **Participants of both the 15-month and 3-year post-COVID studies (N=72)** | **Participants of only the 3-year post-COVID study (N=27)** | **P-value** |
| --- | --- | --- | --- |
| Age, mean (SD) | 55.3 (11.8) | 61.5 (13.4) | 0.09 |
| Male Sex | 41 (56.9%) | 13 (48.2%) | 0.43 |
| Mechanically ventilated, N (%) | 39 (54.2%) | 10 (37.0%) | 0.13 |
| SOFA, mean (SD) | 3.7 (2.9) | 3.6 (2.3) | 0.86 |
| Hospital LOS days, mean (IQR) | 30 (9-48) | 13 (5-34) | 0.06 |
| Ventilator days, median (IQR) | 8 (0-38) | 0 (0-17) | 0.15 |
| Fibrotic ILA | 42 (58.3%) | 17 (63.0%) | 0.68 |
| LTL percentile, mean (SD) | 49.8 (24.3) | 54.1 (28.5) | 0.46 |
| Received Steroids, n (%) | 35 (48.6%) | 9 (33.3%) | 0.17 |
| FVC % pred, mean (SD)* | 86.6 (17.9) | 87.9 (18.1) | 0.75 |
| Reduced FVC* | 21 (29.2%) | 5 (18.5%) | 0.32 |
| DLCO % Predicted, Mean (SD)* | 80.5 (20.5) | 84.65 (19.5) | 0.37 |
| Reduced DLCO* | 32 (44.4%) | 7 (25.9%) | 0.09 |
| 6MWD, % Predicted, mean (SD)† | 74.7 (15.6) | 69.4 (24.3) | 0.21 |
| Reduced 6MWD, N (%)† | 41 (61.2%) | 16 (59.3%) | 0.86 |
| Weak Grip | 25 (34.7%) | 15 (55.6%) | 0.06 |
| Decreased Activity‡ | 3 (4.2%) | 7 (26.0%) | **0.004** |
| Exhaustion | 20 (27.8%) | 11 (40.7%) | 0.22 |
| Slow 4.57 M Walk Speed | 17 (23.6%) | 9 (33.3%) | 0.46 |
| Weight Loss > 10 lbs from Admission | 12 (16.7%) | 6 (22. 2%) | 0.53 |
| Prefrail | 41 (56.9%) | 16 (59.3%) | 0.84 |
| Frail | 6 (8.3%) | 6 (22.2%) | 0.06 |
| Frail or Prefrail | 47 (65.3%) | 22 (81.5%) | 0.12 |

*Data missing from one participant who participated only in the 3-year post-COVID study.

†Data missing from 5 participants who participated in both the 15-month and 3-year post-COVID studies (N=67).

‡Cut off defining decreased activity as previously reported^11^.

**Table S4. Prevalence of HRCT abnormalities 3-years post-COVID-19**

|  | **Total**  **(N=102)** | **Non-Fibrotic Patters**  **(N=40)** | **Fibrotic-like Patterns (N=62)** |
| --- | --- | --- | --- |
| Any Abnormality | 69 (67.6%) |  |  |
| Non-Fibrotic Abnormalities*: |  |  |  |
| Intraparenchymal Opacities, N (%) | 1 (1.0%) | 1 (2.5%) | 0 (0.0%) |
| Ground Glass Opacities, N (%) | 12 (11.8%) | 5 (12.5%) | 7 (11.3%) |
| Nonemphysematous Opacities, N (%) | 3 (2.9%) | 1 (2.5%) | 2 (3.2%) |
| Other Centrilobular Nodules, N (%) | 0 (0.0%) | 0 (0.0%) | 0 (0.0%) |
| Fibrotic Abnormalities*: |  |  |  |
| Reticulations, N (%) | 58 (56.9%) | 0 (0.0%) | 58 (93.6%) |
| Honeycombing, N (%) | 1 (1.0%) | 0 (0.0%) | 1 (1.6%) |
| Traction Bronchiectasis, N (%) | 46 (45.1%) | 0 (0.0%) | 46 (74.2%) |

HRCT: high resolution computed tomography

*Patterns are not mutually exclusive and participants with fibrotic-like patterns can have non-fibrotic patterns.

**Table S5. Semi-quantitative HRCT scores of participants**

|  | **All Scans** | | **Scans with Non-zero Scores** | |
| --- | --- | --- | --- | --- |
| **At 4-months** | **N** | **Median (IQR)** | **N** | **Median (IQR)** |
| Reticulations | 44 | 1.8 (0-5.4) | 31 | 3.6 (1.6-6.0) |
| Traction Bronchiectasis | 44 | 1.0 (0 – 2.0) | 24 | 3.0 (1.5-5.0) |
| Honeycombing | 44 | 0 (0-0) | 1 | 3.4 |
| Ground Glass Opacities | 44 | 0 (0-0.1) | 11 | 2.2 (0.4-7.4) |
| Intraparenchymal Opacities | 44 | 0 (0-0) | 1 | 1.6 |
| Nonemphysematous Cysts | 44 | 0 (0-0) | 1 | 0.2 |
| Centrilobular Nodules | 44 | 0 (0-0) | 0 | NA |
| **At 15-months** | **N** | **Median (IQR)** | **N** | **Median (IQR)** |
| Reticulations | 72 | 0.9 (0 – 4.2) | 46 | 3.0 (1.2-6.6) |
| Traction Bronchiectasis | 72 | 0 (0 – 2.0) | 28 | 2.0 (1.0-4.0) |
| Honeycombing | 72 | 0 (0-0) | 1 | 3.8 |
| Ground Glass Opacities | 72 | 0 (0-0) | 13 | 2.0 (1.2 – 4.0) |
| Intraparenchymal Opacities | 72 | 0 (0-0) | 0 | NA |
| Nonemphysematous Cysts | 72 | 0 (0-0) | 2 | 5.1 (2.4-7.8) |
| Centrilobular Nodules | 72 | 0 (0-0) | 0 | NA |
| **At 3-years** | **N** | **Median (IQR)** | **N** | **Median (IQR)** |
| Reticulations | 102 | 1.4 (0 - 4.2) | 67 | 2.6 (1.4 – 5.8) |
| Traction Bronchiectasis | 102 | 0 (0 – 1.0) | 46 | 1.5 (1.0 – 3.0) |
| Honeycombing | 102 | 0 (0-0) | 1 | 4.8 |
| Ground Glass Opacities | 102 | 0 (0-0) | 14 | 1.8 (0.4 – 3.8) |
| Intraparenchymal Opacities | 102 | 0 (0-0) | 1 | 1.4 |
| Nonemphysematous Cysts | 102 | 0 (0-0) | 4 | 1.9 (0.3 – 5.7) |
| Centrilobular Nodules | 102 | 0 (0-0) | 0 | NA |

HRCT: high resolution computed tomography; NA: not applicable

**Table S6. Spearman correlations between HRCT scores at 3-years post-COVID with measures of pulmonary function, frailty, grip strength and dyspnea**

|  | **FVC Percent Predicted at Year 3 (N=102)** | | | **DLCO Percent Predicted at Year 3 (N=102)** | | | **Six Minute Walk Distance at Year 3 (N=97)** | | |
| --- | --- | --- | --- | --- | --- | --- | --- | --- | --- |
|  | r2 | R | p-value | r2 | R | p-value | r2 | R | p-value |
| **GGO Score** | <0.01 | -0.06 | 0.58 | 0.01 | 0.07 | 0.50 | 0.01 | 0.11 | 0.28 |
| **Reticulation Score** | <0.01 | -0.02 | 0.83 | 0.09 | -0.31 | **0.002** | 0.01 | -0.09 | 0.38 |
| **Traction Bronchiectasis Score** | 0.01 | -0.08 | 0.45 | 0.04 | -0.21 | **0.04** | <0.01 | 0.01 | 0.90 |
|  | **Frailty Score at Year 3 (N=97)** | | | **Grip Strength at Year 3 (N=102)** | | | **UCSD SOBQ at Year 3 (N=102)** | | |
|  | r2 | R | p-value | r2 | R | p-value | r2 | R | p-value |
| **GGO Score** | <0.01 | -0.06 | 0.58 | <0.01 | 0.02 | 0.85 | 0.01 | 0.10 | 0.30 |
| **Reticulation Score** | 0.04 | 0.19 | 0.06 | 0.01 | 0.08 | 0.43 | 0.06 | 0.24 | **0.02** |
| **Traction Bronchiectasis Score** | 0.02 | 0.12 | 0.22 | 0.05 | 0.22 | **0.03** | 0.06 | 0.25 | **0.01** |

Abbreviations: FVC, forced vital capacity; FEV1, forced expiratory volume in 1 second; DLCO, diffusion capacity for carbon monoxide; 6MWD, six-minute walk distance SOBQ, shortness of breath questionnaire; GGO, ground glass opacities

**Table S7. Pairwise comparisons of participants with HRCT scans with non-zero HRCT scores**

|  | **Ground Glass Opacities** | | | **Reticulations** | | | **Traction Bronchiectasis** | | |
| --- | --- | --- | --- | --- | --- | --- | --- | --- | --- |
|  | N | Score, Median (IQR) | P-value | N | Score, Median (IQR) | P-value | N | Score, Median (IQR) | P-value |
| Individuals with non-zero scores at 4-mo and imaging at two time points: |  |  |  |  |  |  |  |  |  |
| At 4-mo post-COVID | 10 | 2.9 (1-7.4) | **0.02** | 28 | 3.4 (1.6-5.9) | **<0.001** | 23 | 3 (1-5) | **0.001** |
| At 15-mo post-COVID | 10 | 2.8 (0.2-4.2) |  | 28 | 2.3 (1.1-4.9) |  | 23 | 2 (1-5) |  |
| At 4-mo post-COVID | 11 | 2.2 (0.4-7.4) | **0.04** | 31 | 3.6 (1.6- 6.0) | **<0.001** | 24 | 3 (1.5-5) | **0.002** |
| At 3-yr post-COVID | 11 | 1.8 (0.2-5.0) |  | 31 | 1.8 (1.0-5.2) |  | 24 | 2.5 (1-3.5) |  |
| At 15-yr post-COVID | 10 | 2.8 (0.2-4.2) | 0.68 | 28 | 2.3 (1.1-4.9) | **0.03** | 23 | 2 (1-5) | 0.92 |
| At 3-yr post-COVID | 10 | 2.1 (0.2-5) |  | 28 | 2.1 (1.1-4.7) |  | 23 | 2 (1-4) |  |
| Individuals with non-zero scores at 15-mo and imaging at two time points |  |  |  |  |  |  |  |  |  |
| At 4-mo post-COVID | 8 | 4.6 (1.6- 7.9) | **0.04** | 27 | 3.6 (1.6-6.0) | **0.0004** | 18 | 4 (2-5) | **0.02** |
| At 15-mo post-COVID | 8 | 3.8 (1.2- 4.2) |  | 27 | 2.6 (1.2-5.0) |  | 18 | 3 (2-5) |  |
| At 4-mo post-COVID | 8 | 4.6 (0.4-7.9) | 0.079 | 27 | 3.6 (1.6-6.0) | **<0.001** | 18 | 4 (2-5) | **0.006** |
| At 3-yr post-COVID | 8 | 3.1 (1-5.1) |  | 27 | 2.4 (1.2-5.2) |  | 18 | 3 (2-4) |  |
| At 15-mo post-COVID | 13 | 2 (1.2-4) | 0.38 | 46 | 3.0 (1.2-6.6) | **0.005** | 28 | 2 (1-4) | 0.06 |
| At 3-yr post-COVID | 13 | 1.8 (0.2-3.8) |  | 46 | 2.6 (1.2-5.8) |  | 28 | 2 (1-3.5) |  |
| Individuals with non-zero scores at 3-yr and imaging at two time points: |  |  |  |  |  |  |  |  |  |
| At 4-mo post-COVID | 10 | 2.9 (0.4-7.4) | **0.02** | 28 | 3.4 (1.6-5.9) | **0.0002** | 24 | 3 (1-5) | **0.001** |
| At 15-mo post-COVID | 10 | 2.8 (0.2-4.2) |  | 28 | 2.3 (1.1-4.9) |  | 24 | 2 (0.5-4) |  |
| At 4-mo post-COVID | 10 | 2.9 (0.4-7.4) | 0.13 | 31 | 3.6 (1.6-6.0) | **<0.001** | 25 | 3 (1-5) | **0.006** |
| At 3-yr post-COVID | 10 | 2.1 (0.4-5) |  | 31 | 1.8 (1.0-5.2) |  | 25 | 2 (1-3) |  |
| At 15-mo post-COVID | 14 | 1.9 (0.2-4.0) | 0.39 | 46 | 3.0 (1.0-6.6) | **0.02** | 36 | 2.0 (1-3) | 0.91 |
| At 3-yr post-COVID | 14 | 1.8 (0.4-3.8) |  | 46 | 2.6 (1.2-5.8) |  | 36 | 1.0 (1-3) |  |

HRCT: high resolution computed tomography

**Table S8. Qualitative assessment of HRCT changes after COVID-19**

|  | **Between 4-mo and 15-mo**  **(N=43)** | **Between 15-mo and 3-yr**  **(N=77)** | **Between 4-mo and 3-yr**  **(N=56)** |
| --- | --- | --- | --- |
| Improvement, N (%) | 10 (19.6%) | 7 (9.1%) | 16 (28.6%) |
| Stability, N (%) | 40 (78.4%) | 70 (90.2%) | 40 (71.4%) |
| Worsening, N (%) | 0 (0%) | 0 (0%) | 0 (0%) |

HRCT: high resolution computed tomography.

**­**

**Table S9. Quantification of imaging findings, pulmonary function, indices of frailty and symptom scores for all participants and for the subset of participants with follow-up at both 15-months and 3-years after COVID-19**

| **Abnormality** | **All Participants at 15-month, (N=104)** | **All Participants at 3-years, (N=102)** | **Subset with follow-up at both time points, at 15-month, (N=72)** | **Subset with follow-up at both time points, at 3-years, (N=72)** | **P-Value** |
| --- | --- | --- | --- | --- | --- |
| Imaging Findings |  |  |  |  |  |
| Any imaging Abnormality, N (%) | 67 (64.0%) | 69 (67.6%) | 45 (62.5%) | 48 (66.7%) | 0.12 |
| Any Fibrotic-like Abnormality, N (%) | 67 (64.0%) | 62 (60.8%) | 40 (55.6%) | 42 (58.3%) | 0.38 |
| Air Trapping, N (%)* | 43 (68.2%) | 76 (80.8%) | 18 (48.6%) | 31 (88.7%) | **<0.001** |
| Pulmonary Function |  |  |  |  |  |
| FVC % pred, mean (SD)† | 90.4 (17.2) | 86.9 (17.7) | 88.7 (16.8) | 86.6 (17.9) | 0.13 |
| Reduced FVC, N (%)‡ | 18 (17.0%) | 27 (26.5%) | 14 (19.4%) | 21 (29.2%) | **0.02** |
| Reduced FEV1/FVC | 5 (4.0%) | 1 (1.0%) | 3 (3.2%) | 1 (1.4%) | 0.50 |
| DLCO % Predicted, mean (SD)‡ | 80.9 (19.1) | 81.2 (20.2) | 81.3 (18.8) | 80.5 (20.5) | 0.71 |
| Reduced DLCO, N(%)‡ | 40 (39.0%) | 41 (40.2%) | 27 (38.0%) | 32 (44.4%) | 0.47 |
| 6MWD, median (IQR)§ | 399 (344-469) | 375 (327-417) | 369 (309-450) | 384 (346-424) | 0.65 |
| 6MWD, Percent Predicted§ | 80.9 (19.1) | 72.8 (18.4) | 73.2 (20.7) | 74.7 (15.6) | 0.60 |
| Reduced 6MWD§ | 58 (58.0%) | 60 (61.9%) | 45 (62.3%) | 41 (61.2%) | 0.80 |
| Indices of Frailty |  |  |  |  |  |
| Weak Grip | 46 (44.2%) | 41 (40.2%) | 33 (45.8%) | 25 (34.7%) | 0.06 |
| Decreased Activity | 1 (1.0%) | 10 (9.8%) | 0 (0%) | 3 (4.2%) | 0.25 |
| Exhaustion | 31 (29.8%) | 32 (31.4%) | 27 (37.5%) | 20 (27.8%) | 0.17 |
| Slow 4.57 m Walk Speed | 12 (11.5%) | 29 (28.4%) | 9 (12.5%) | 17 (23.6%) | 0.08 |
| Weight Loss > 10 lbs | 10 (9.6%) | 19 (18.6%) | 9 (12.5%) | 12 (16.7%) | 0.55 |
| Prefrail (1-2 Criteria) | 56 (53.8%) | 60 (58.8%) | 46 (63.9%) | 41 (56.9%) | 0.40 |
| Frail (>3 Criteria) | 7 (6.7%) | 12 (11.8%) | 4 (5.6%) | 6 (8.3%) | 0.73 |
| Symptom Scores |  |  |  |  |  |
| MMRC Dyspnea Score, median ( IQR) | 1 (0-2) | 1 (0-2) | 1 (0-2) | 1 (0-2) | 0.84 |
| MMRC Dyspnea Score ≥3 | 13 (12.4%) | 15 (14.2%) | 10 (13.9%) | 10 (13.9%) | 1.0 |
| UCSD SOBQ Score, median (IQR) | 30 (15-51) | 30 (9-55) | 32.5 (20-57) | 30 (10.5-56.5) | **0.02** |
| Cough VAS score, median (IQR) | 0 (0-20) | 16.5 (0-20) | 0 (0-27.5) | 28.5 (5.5-50) | **<0.001** |
| Cough Severity ≥30 | 18 (17.3%) | 38 (37.2%) | 18 (25.0%) | 35 (48.6%) | **<0.001** |

*The presence of air trapping was determined by consensus of two thoracic radiologists. Due to the limited quality of some of the expiratory scans, air trapping could only be assessed for 63 participants at 15-month and 94 participants at 3-years (N=35 for the subset with both time points).

†Data missing for one individual at 3-year post-COVID

‡Data missing for 2 individuals (N=102 at 15-month, N=101 at 3-years, N=71 for subset with follow up at both time points at 15-month)

§Data missing for 6 individuals (N=100 at 15-month, N=97 at 3-years, N=70 for subset with follow up at both time points, at 15-month, N=67 for subset with follow up at both time points, at 3-years)

**Table S10. Associations with air trapping noted on expiratory imaging studies at 3-year post-COVID-19**

|  | **Air Trapping on 3-year Imaging Study**  **N=76 (81%)*** | **No Air Trapping on 3-year Imaging Study**  **N=18 (19%)*** | **P- Value** |
| --- | --- | --- | --- |
| Fibrotic ILA, N (%) | 41 (54.0%) | 15 (83.3%) | **0.02** |
| Reticulation score, Median (IQR) | 0.7 (0-3.7) | 2.5 (1.6-4.6) | 0.057 |
| Cough VAS ≥30 | 28 (46.8%) | 8 (44.1%) | 0.55 |
| Cough VAS, Median (IQR) | 15.5 (0-49.5) | 26.5 (4-70) | 0.18 |
| Spring Visit, N (%) | 38 (50%) | 9 (50%) | 1 |
| Self-reported Asthma, N (%) | 21 (27.63%) | 1 (5.5%) | **0.02** |
| Self-reported COPD, N (%) | 4 (5.3%) | 1 (5.5%) | 1 |
| FVC % Pred, Mean (SD)† | 87.6 (17.75) | 85.72 (16.63) | 0.38 |
| FEV1/FVC % Predicted, Mean (SD)† | 101.51 (6.9) | 102.22 (12.42) | 0.71 |
| Low FEV1/FVC, N (%) | 0 (0%) | 1 (5.5%) | 0.19 |

* The presence of air trapping was determined by consensus of two thoracic radiologists. Due to the limited quality of some of the expiratory scans, air trapping could only be assessed for 94 participants at 3-years.

†Data missing for 1 individual (N=75 for those with air trapping)

**Table S11. Number of participants with and without fibrotic-like abnormalities on their 3-year HRCT chest scans who have self-reported post-COVID symptoms**

|  | **Total** | **Normal or Non-Fibrotic Abnormalities** | **Fibrotic ILA** | **P-Value** |
| --- | --- | --- | --- | --- |
| N | 102 | 40 | 62 |  |
| Fatigue | 42 (41.2%) | 12 (30%) | 30 (48.4%) | 0.07 |
| Weakness | 37 (36.3%) | 12 (30%) | 25 (40.3%) | 0.29 |
| Night Sweats | 19 (18.6%) | 6 (15%) | 13 (21%) | 0.46 |
| Fever or Chills | 7 (6.9%) | 3 (7.5%) | 4 (6.5%) | 0.84 |
| Rash | 8 (7.8%) | 1 (2.5%) | 7 (11.3%) | 0.11 |
| Muscle Pain, Cramps, Body Aches | 35 (34.3%) | 13 (32.5%) | 22 (35.5%) | 0.76 |
| Tremor or Shakiness | 14 (13.7%) | 4 (10%) | 10 (16.1%) | 0.38 |
| Numbness or Tingling of Hands and Feet | 27 (26.5%) | 9 (22.5%) | 18 (29%) | 0.47 |
| Joint Pain | 33 (32.4%) | 12 (30%) | 21 (33.9%) | 0.69 |
| Weight Gain | 8 (7.8%) | 3 (7.5%) | 5 (8.1%) | 0.92 |
| Hair Loss | 25 (24.5%) | 13 (32.5%) | 12 (19.4%) | 0.13 |
| Headache | 19 (18.6%) | 8 (20%) | 11 (17.7%) | 0.78 |
| Difficulty Focusing | 24 (23.5%) | 11 (27.5%) | 13 (21%) | 0.45 |
| Confusion | 20 (19.6%) | 8 (20%) | 12 (19.4%) | 0.94 |
| Memory Problems | 27 (26.5%) | 12 (30%) | 15 (24.2%) | 0.52 |
| Sadness | 16 (15.7%) | 5 (12.5%) | 11 (17.7%) | 0.48 |
| Anxiety | 22 (21.6%) | 9 (22.5%) | 13 (21%) | 0.86 |
| Personality Changes | 13 (12.7%) | 6 (15%) | 7 (11.3%) | 0.59 |
| Sleeping More than Normal | 13 (12.7%) | 4 (10%) | 9 (14.5%) | 0.51 |
| Difficulty Sleeping | 37 (36.3%) | 12 (30%) | 25 (40.3%) | 0.29 |
| Dizziness | 14 (13.7%) | 4 (10%) | 10 (16.1%) | 0.38 |
| Tinnitus | 9 (8.8%) | 2 (5%) | 7 (11.3%) | 0.28 |
| Clogged Ears | 8 (7.8%) | 4 (10%) | 4 (6.5%) | 0.52 |
| Blurry Vision | 18 (17.6%) | 6 (15%) | 12 (19.4%) | 0.58 |
| Loss or Change in Smell | 11 (10.8%) | 5 (12.5%) | 6 (9.7%) | 0.66 |
| Constant Thirst | 15 (14.7%) | 6 (15%) | 9 (14.5%) | 0.95 |
| Nasal Congestion or Runny Nose | 22 (21.6%) | 8 (20%) | 14 (22.6%) | 0.76 |
| Cough | 17 (16.7%) | 4 (10%) | 13 (21%) | 0.15 |
| Sore Throat | 8 (7.8%) | 4 (10%) | 4 (6.5%) | 0.52 |
| Shortness of Breath at Rest | 19 (18.6%) | 7 (17.5%) | 12 (19.4%) | 0.82 |
| Shortness of Breath with Exertion | 31 (30.4%) | 12 (30%) | 19 (30.6%) | 0.95 |
| Chest Pain | 9 (8.8%) | 3 (7.5%) | 6 (9.7%) | 0.71 |
| Racing Heartbeat | 10 (9.8%) | 6 (15%) | 4 (6.5%) | 0.16 |
| Gastric Reflux | 14 (13.7%) | 6 (15%) | 8 (12.9%) | 0.77 |
| Abdominal Pain | 12 (11.8%) | 6 (15%) | 8 (12.9%) | 0.66 |
| Back Pain | 32 (31.4%) | 11 (27.5%) | 21 (33.9%) | 0.50 |
| Nausea or Vomiting | 6 (5.9%) | 1 (2.5%) | 5 (8.1%) | 0.25 |
| Diarrhea | 5 (4.9%) | 1 (2.5%) | 4 (6.5%) | 0.37 |
| Change in Urinary Frequency | 17 (16.7%) | 5 (12.5%) | 12 (19.4%) | 0.37 |
| Constipation | 18 (17.6%) | 7 (17.5%) | 11 (17.7%) | 0.98 |
| At Least One Symptom Moderately bothersome or greater | 83 (81.4%) | 29 (72.5%) | 54 (87.1 %) | 0.06 |

**Table S12. HRCT Scores of five participants who underwent bronchoscopy with transbronchial biopsy**

|  | **4-months post-COVID** | **15-month post-COVID** | **3-years post-COVID** |
| --- | --- | --- | --- |
| **Participant #1:** |  |  |  |
| Reticulations | 7.8 | 6.8 | 7 |
| Traction Bronchiectasis | 4 | 2 | 3 |
| Honeycombing | 0 | 0 | 0 |
| Intraparenchymal Opacities | 0 | 0 | 0 |
| Ground Glass Opacities | 0 | 0 | 0 |
| Nonemphysematous Cysts | 0 | 0 | 0 |
| Diffuse Centrilobular Nodules | 0 | 0 | 0 |
| **Participant #2:** |  |  |  |
| Reticulations | 8.4 | 7.8 | 6.8 |
| Traction Bronchiectasis | 5 | 3 | 4 |
| Honeycombing | 0 | 0 | 0 |
| Intraparenchymal Opacities | 0 | 0 | 0 |
| Ground Glass Opacities | 0 | 0 | 0 |
| Nonemphysematous Cysts | 0 | 0 | 0 |
| Diffuse Centrilobular Nodules | 0 | 0 | 0 |
| **Participant #3:** |  |  |  |
| Reticulations | NA | 7.4 | 7 |
| Traction Bronchiectasis | NA | 1 | 1 |
| Honeycombing | NA | 0 | 0 |
| Intraparenchymal Opacities | NA | 0 | 0 |
| Ground Glass Opacities | NA | 0 | 0 |
| Nonemphysematous Cysts | NA | 0 | 0 |
| Diffuse Centrilobular Nodules | NA | 0 | 0 |
| **Participant #4:** |  |  |  |
| Reticulations | NA | 7.2 | 7.2 |
| Traction Bronchiectasis | NA | 0 | 0 |
| Honeycombing | NA | 0 | 0 |
| Intraparenchymal Opacities | NA | 0 | 0 |
| Ground Glass Opacities | NA | 0 | 0 |
| Nonemphysematous Cysts | NA | 0 | 0 |
| Diffuse Centrilobular Nodules | NA | 0 | 0 |
| **Participant #5:** |  |  |  |
| Reticulations | 8 | NA | 7 |
| Traction Bronchiectasis | 4 | NA | 3 |
| Honeycombing | 0 | NA | 0 |
| Intraparenchymal Opacities | 0 | NA | 0 |
| Ground Glass Opacities | 0 | NA | 0 |
| Nonemphysematous Cysts | 0 | NA | 0 |
| Diffuse Centrilobular Nodules | 0 | NA | 0 |

HRCT: high resolution computed tomography; NA: not available

**Figure S1. Recruitment Flow Diagram**

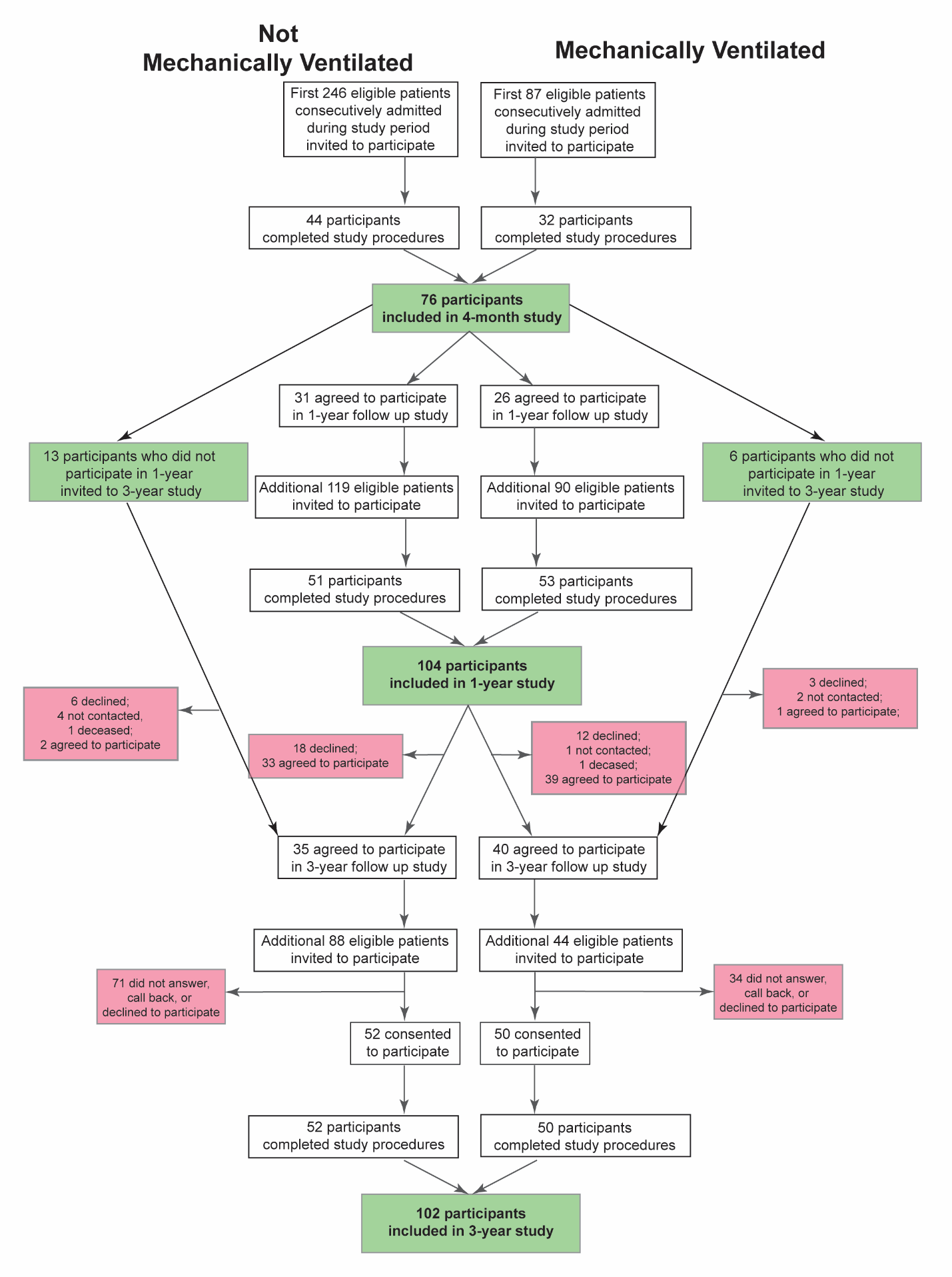

**Figure S2. HRCT Findings of Five Participants who Underwent Bronchoscopy with Transbronchial Lung Biopsy**

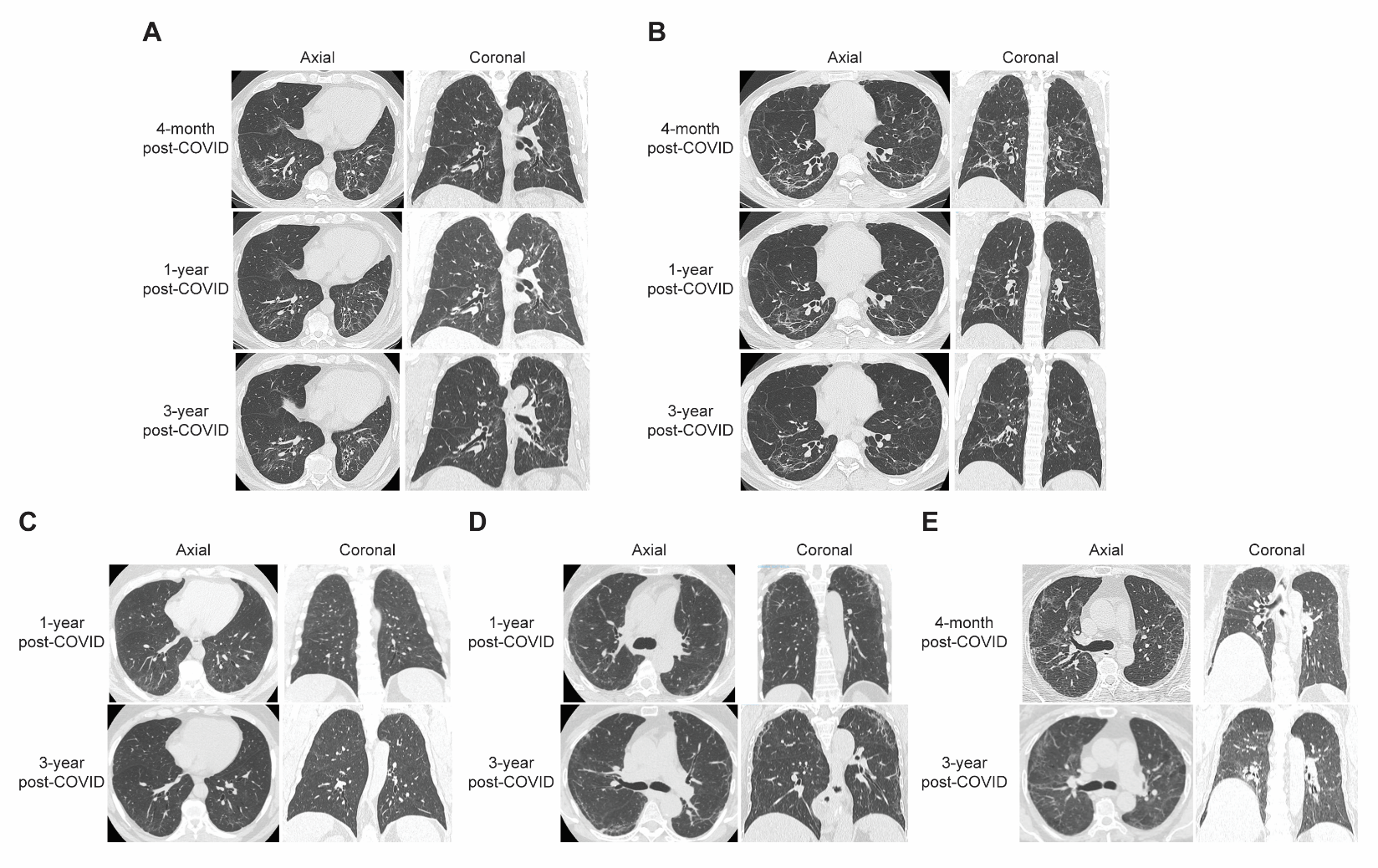

**­REFERNCES**
